# Behaviour of smokers and their influencers in the UK during COVID-19 pandemic

**DOI:** 10.1101/2021.05.25.21257716

**Authors:** Sudhanshu Patwardhan, Claudia Trainer

## Abstract

**Purpose:** The COVID-19 pandemic has resulted in unprecedented circumstances and changes in behaviour. This research sought to better understand the impact of the COVID-19 pandemic, and lockdown, on smoking behaviour in the UK from the perspectives of consumers (current and former smokers) and some of their smoking-related behaviour-influencers.

**Design/methodology/approach:** This research project encompassed two surveys, one for current and former smokers (Consumers) and one for those individuals in professions with the potential to influence smoking behaviours (Influencers). Both surveys were conducted online and were infield for approximately two weeks during UK’s first COVID-19 lockdown. Because of the unprecedented times the society was experiencing, several questions relating directly to COVID-19 were added to the survey and this paper is based only on findings only from those questions and not the whole project. The results were analysed descriptively.

**Findings:** A total of 954 consumers and 1027 influencers participated in the surveys. Increased smoking was reported by 67% of the consumers mainly due to stress and boredom arising out of COVID-19 lockdown. Consumers under 45 years of age, those in professional and managerial occupations, and among dual users reported increased smoking in lockdown. The COVID-19 situation changed the plans to quit smoking in 36% of consumers, with only 6% deciding to quit. Only 40% of healthcare professionals (HCPs) documented patient smoking status in over half their interactions.

**Originality/ value:** This research among current and former smokers and their influencers highlights important changes in behaviour during the COVID-19 times and underscores urgent measures to be taken by HCPs and policymakers for staying on course of achieving smokefree goals despite challenges posed by COVID-19.

## Background

The COVID-19 pandemic has been high on the public agenda for much of 2020 and into 2021. Early evidence from China suggested that smokers were 14 times more likely to develop severe complications from COVID-19 (1), a message that was promoted by Public Health England (2). Initiatives such as ‘Quit for COVID’ from Smokefree Action were launched during this time, attempting to use this pivotal time to encourage positive change (3). According to research by Action on Smoking and Health (ASH) and University College London, over one million smokers in the UK quit smoking and a further 440,000 attempted to do so, since the beginning of the pandemic (4). However, this conclusion was based on a survey done by YouGov on a small sample population. Concerns around increased risk of relapse and an increase in smoking rates as a result of COVID-19 measures were voiced by smoking cessation practitioners (5, 6).

Over the last decade, the UK has made good progress in reducing the prevalence of smoking in adults, with Public Health England (PHE) reporting a fall from 19.8% in 2011 to 14.1% in 2019 (7, 8). However, there is some way to go to achieve the stated goal of a ‘smokefree’ England (<5% prevalence) by 2030 (9, 10). At present, smoking remains the leading cause of preventable death and disease in the UK, with approximately 1 in 6 deaths being attributable to smoking (11).

Office for National Statistics (ONS) data suggests that the majority (62.5%) of smokers in the UK who have ever smoked had quit in 2019, while of those who currently smoke 52.7% intended to quit with 21.1% intending to quit in the next three months (8). Knowing that the UK already has some of the strictest tobacco control measures in the world, it is crucial to understand the barriers that the remaining smokers in the UK are facing to quit smoking sustainably. To understand these barriers, we conducted a qualitative and quantitative research study on a convenience sample of the UK’s remaining smokers and their influencers, i.e. professionals who might have a positive influence on smokers’ quitting journey. The overarching aim was that these findings will help to design targeted interventions that, if implemented, could accelerate the UK’s journey to becoming ‘smokefree’.

The qualitative phase of our research helped us better understand the various stakeholder groups and their role in the influencer ecosystem. The quantitative phase originally encompassed two surveys, one for current and former smokers (referred to as ‘consumers’ throughout) and one for individuals in some of the professions with the potential to influence smoking (referred to as ‘influencers’ throughout). In addition, beauticians, social workers and barbers/hairdressers were included in the influencer group. While not traditional influencers in this space, in times when national healthcare resources are limited, we wanted to explore if these professions, where there are often repeated encounters with the same individuals, could have a potential to convey healthy lifestyle messages.

Both surveys were conducted online and were infield for approximately two weeks from the 15th of May 2020 – a time when the UK was in its first lockdown.

Given the impact of the COVID-19 pandemic on peoples’ lives, the scope of the planned survey was expanded to incorporate this. Within these surveys, several questions relating directly to COVID-19 were included which form the basis of this paper. The overall qualitative research and remaining quantitative survey findings will be reported elsewhere (manuscripts in various stages of preparation).

The purpose of this paper is to better understand and communicate the impact of the COVID-19 pandemic, and lockdown, on current and former smokers in the UK.

## Methods

### Participants

A total of 1027 influencers were identified by their reported profession and grouped, as shown in Table 1, to aid analysis.

**Table 1:** Influencers by profession and grouping

| Profession | Number of influencer respondents | Categorisation |
| --- | --- | --- |
| General Practitioner | 101 | HCP |
| General Practice Surgery Practice Nurse | 51 | HCP |
| Midwife | 100 | HCP |
| Community Mental Health Nurse | 100 | HCP |
| Dentist | 100 | HCP |
| Pharmacist | 100 | HCP |
| Respiratory Specialist | 75 | HCP |
| Stop Smoking Advisor (SSA) | 50 | HCP |
| Social Worker | 100 | Non-HCP |
| Hairdresser/barber | 100 | Non-HCP |
| Beautician | 100 | Non-HCP |
| Vape Shop Employee/Owner | 50 | Non-HCP |
| **Total** | **1,027** |  |

Notes:
HCP = health care professional

A total of 954 consumers participated in the study. For the purposes of this report, consumers were split into smoker types using their reported smoking and vaping status as shown in Table 2.

**Table 2:**
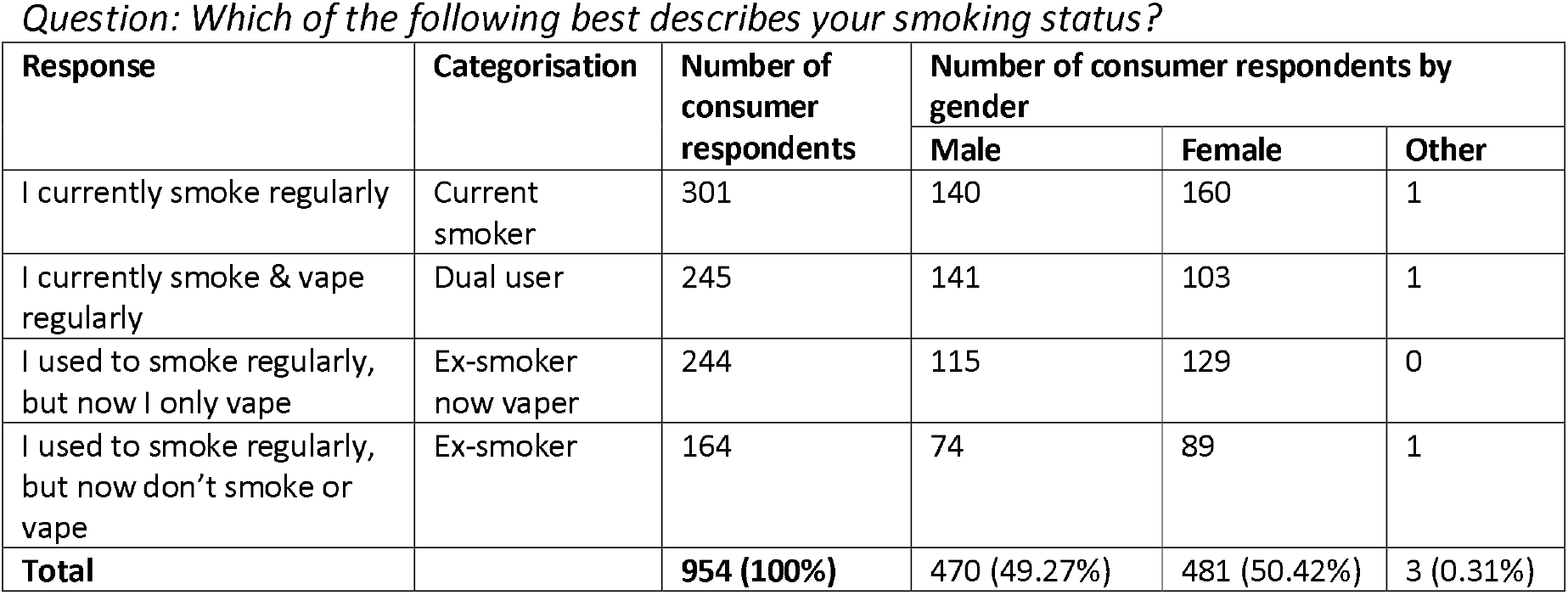
Consumer smoker type definitions and gender split Question: Which of the following best describes your smoking status?

| Response | Categorisation | Number of consumer respondents | Number of consumer respondents by gender |  |  |
| --- | --- | --- | --- | --- | --- |
|  |  |  | Male | Female | Other |
| I currently smoke regularly | Current smoker | 301 | 140 | 160 | 1 |
| I currently smoke & vape regularly | Dual user | 245 | 141 | 103 | 1 |
| I used to smoke regularly, but now I only vape | Ex-smoker now vaper | 244 | 115 | 129 | 0 |
| I used to smoke regularly, but now don't smoke or vape | Ex-smoker | 164 | 74 | 89 | 1 |
| **Total** |  | **954 (100%)** | **470 (49.27%)** | **481 (50.42%)** | **3 (0.31%)** |

**Table 3:**
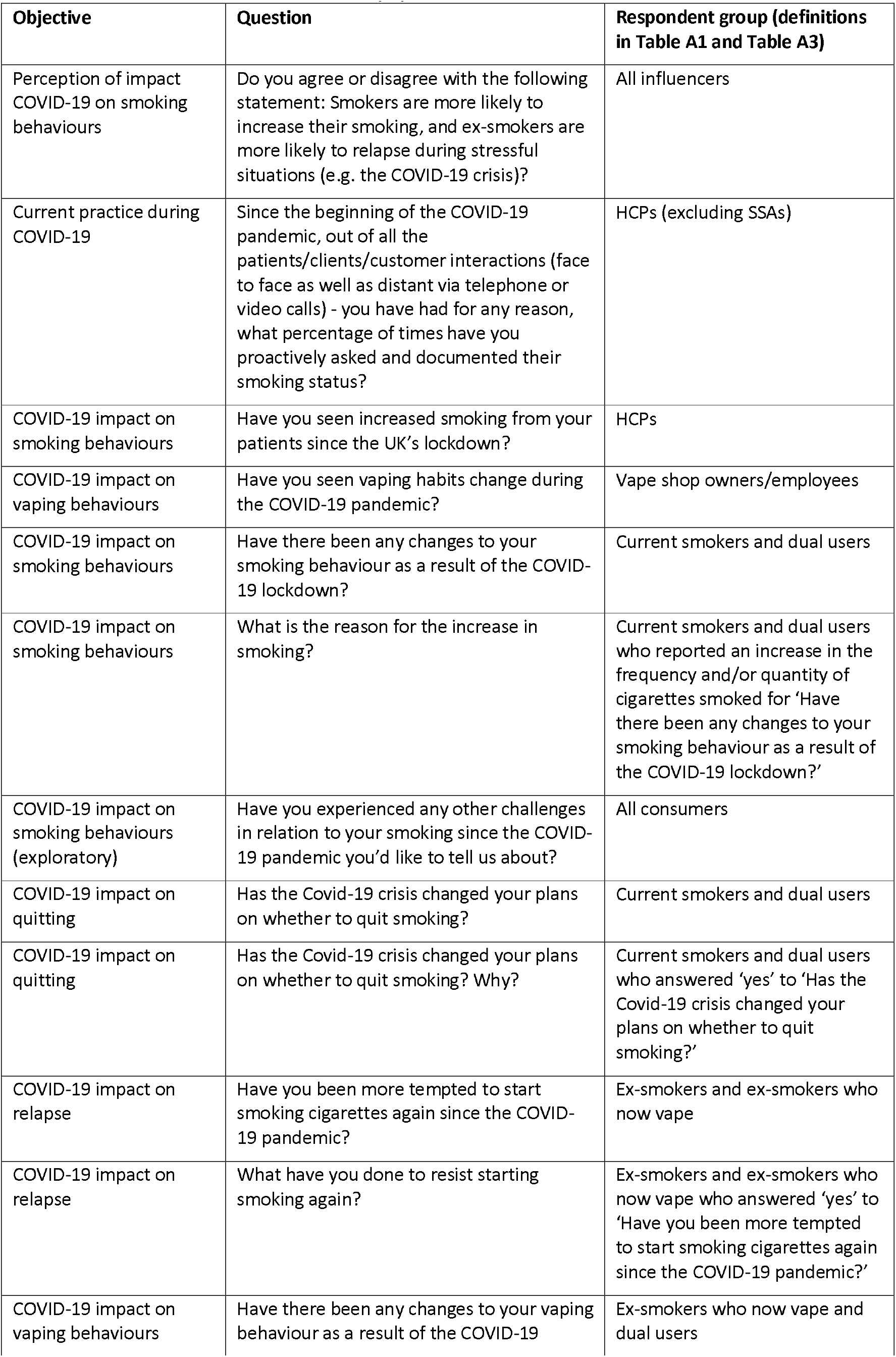
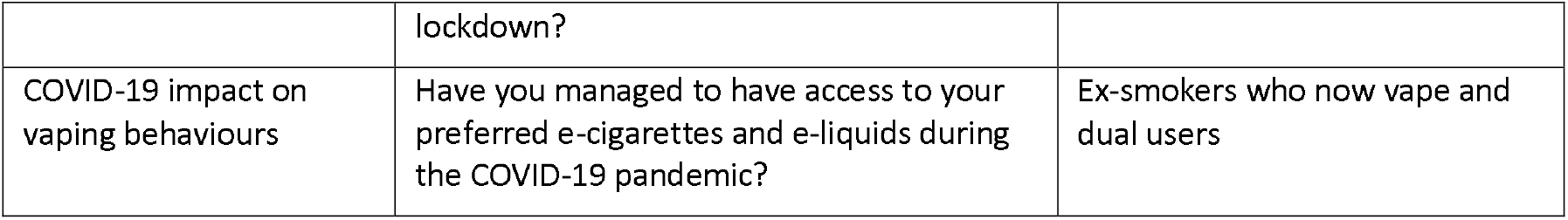
List of Covid-19 related survey questions

Consumers were allocated a social grade (A, B, C1, C2, D, E) based on their profession:

|  |  |
| --- | --- |
| **GRADE A** | High managerial, administrative or professional - e.g. doctor, lawyer, company director (50+ people), judge, surgeon, school headmaster etc. (n=82) |
| **GRADE B** | Intermediate managerial, administrative or professional - e.g. school teacher, office manager, junior doctor, bank manager, police inspector, accountant etc. (n=227) |
| **GRADE C1** | Supervisor, clerical, junior managerial, administrative or professional - e.g. policeman, nurse, secretary, clerk, self-employed (5+ people), students etc. (n=256) |
| **GRADE C2** | Skilled manual worker - e.g. mechanic, plumber, electrician, lorry driver, train driver etc. (n=103) |
| **GRADE D** | Semi-skilled or unskilled manual worker - e.g. baggage handler, waiter, factory worker, receptionist, labourer, gardener etc. (n=88) |
| **GRADE E** | Housewife/Househusband, Retired and living on state pension, Unemployed, and any other consumers not covered under above categories (n=198) |

### Data collection

An online survey was conducted by Atomik Research among 954 consenting adults aged 18+ from the UK. The research fieldwork took place on 19-27 May 2020. Separately, an online survey was conducted by Atomik Research among 1,027 consenting healthcare specialists and other professionals.

During this time, the UK was under lockdown due to COVID-19 and had been since late March, although specific rules varied between nations. However, throughout the UK, lockdown at this time meant that citizens were advised to work from home wherever possible and non-essential businesses were closed. These ‘non-essential’ businesses included dental surgeries and brick and mortar vape shops.

Atomik Research is an independent creative market research agency that employs MRS-certified researchers and abides by the MRS code. Respondents’ participation in this study was anonymous, voluntary and incentivised. For both surveys, respondents were directed to questions based on their previous answers and categorisation, therefore the number of respondents varied per question and is often less than the total number of participants.

### Data analysis

As this study was largely descriptive, no power analysis was conducted. The purpose of this study was to better understand the perceptions and experiences of smoking cessation from multiple perspectives as opposed to testing specific hypotheses. Results are largely presented as averages and percentages. For qualitative results, the software programme NVivo was used to record and structure key themes.

## Results

The following section summarises the results under the following sub-sections:

- Influencer perceptions of the impact of stressful situations (such as COVID-19) on increased smoking and relapse
- The extent of documentation of smoking status during COVID-19 by HCPs (excluding SSAs)
- HCP perception of changes to smoking behaviours due to COVID-19
- Self-reported changes in consumer smoking behaviours due to COVID-19
- Impact of COVID-19 on consumer quit intentions
- Impact of COVID-19 on the temptation to relapse
- Self-reported changes in consumer vaping behaviours due to COVID-19
- Other self-reported challenges faced by smokers during COVID-19

### Influencer perceptions of the impact of stressful situations (such as COVID-19) on increased smoking and relapse

Overall, the majority of both HCPs (85%) and non-HCPs (82%) agreed or strongly agreed that smokers were more likely to increase their smoking and ex-smokers were more likely to relapse during COVID-19 (Figure 1).

**Figure 1.**
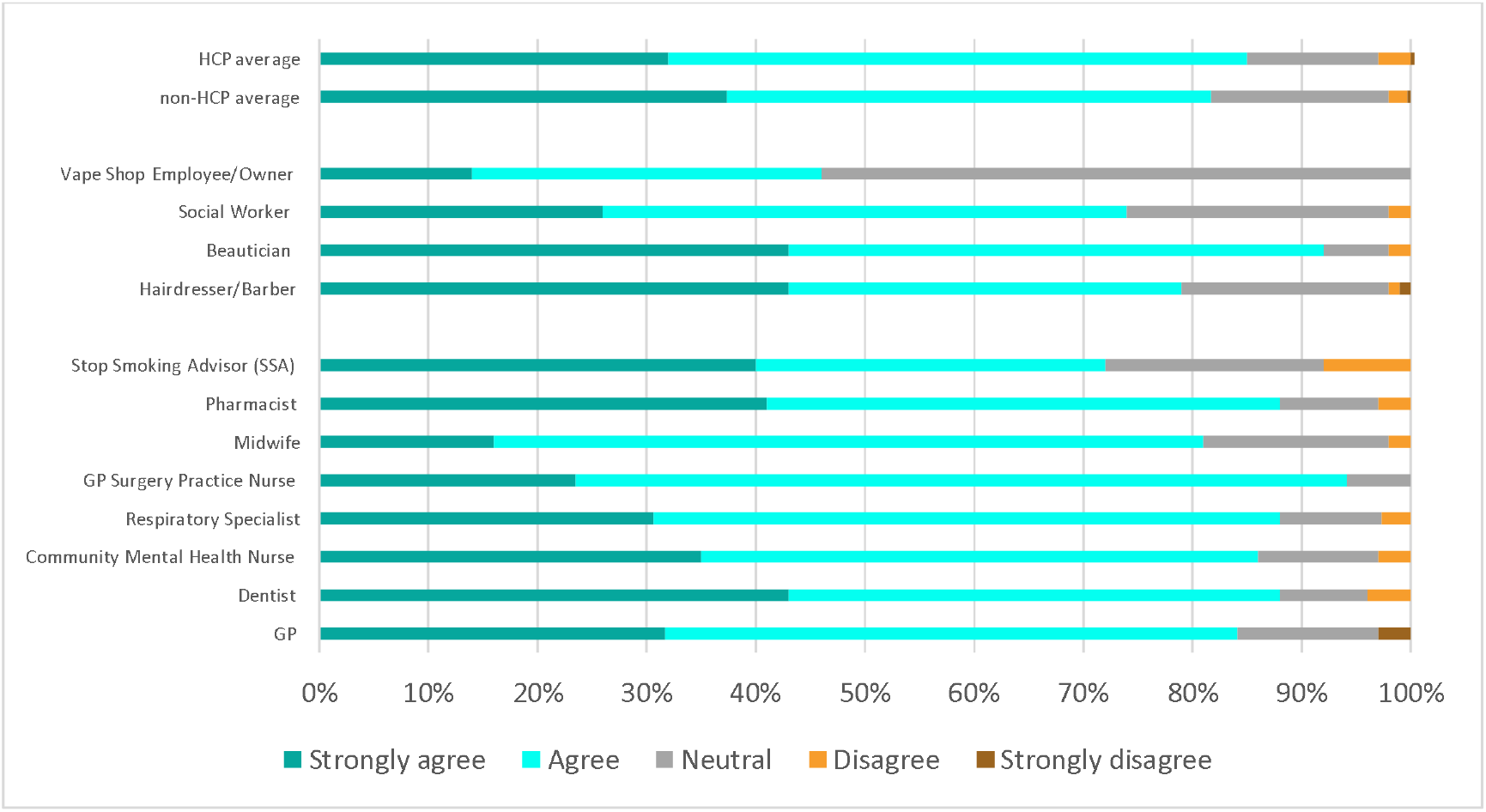
Influencer perception about smoking behaviour during COVID-19 pandemic Question: Do you agree or disagree with the following statement: Smokers are more likely to increase their smoking, and ex-smokers are more likely to relapse during stressful situations (e.g. the COVID-19 crisis)?, N=927 (627 HCPs, 300 Non-HCPs).

### The extent of documentation of smoking status during COVID-19 by HCPs (excluding SSAs)

HCPs, excluding SSAs, were asked in what proportion of interactions (in-person, or distant) with patients they proactively asked and documented their smoking status: 60% of HCPs reported doing so in half or less than half of their interactions (Figure 2).

**Figure 2.**
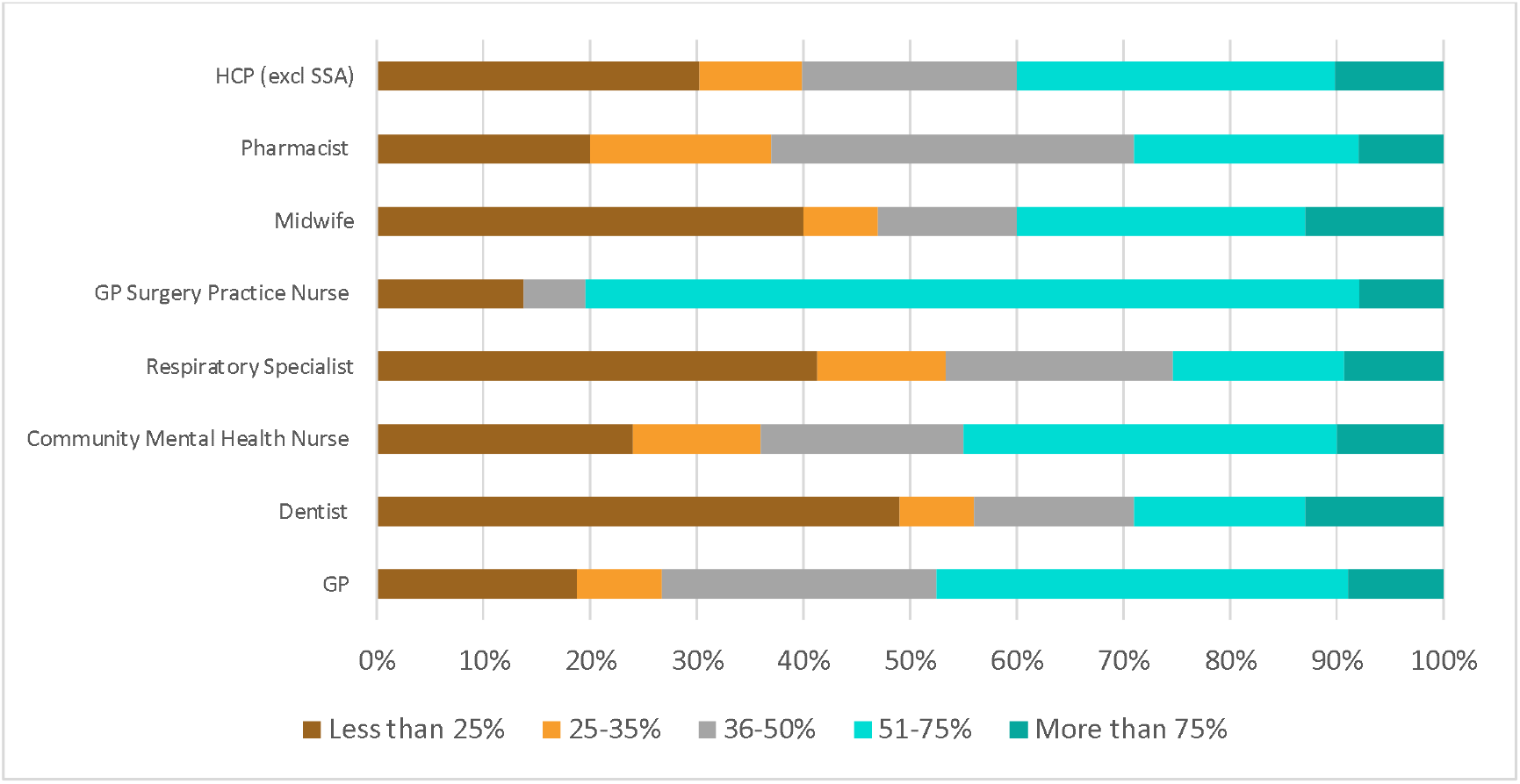
Frequency of requesting and documenting patient smoking status amongst HCPs (excluding SSAs) during COVID-19 Question: Since the beginning of the COVID pandemic, out of all the patients/clients/customer interactions (face to face as well as distant via telephone or video calls) - you have had for any reason, what percentage of times have you proactively asked and documented their smoking status?, N=627 HCP (excluding SSAs).

### HCP perception of changes to smoking behaviours due to COVID-19

A total of 39% of HCPs were unsure as to whether their patients had increased their smoking due to less patient contact or because they didn’t ask all patients (Figure 3), despite their recognition of the increased risk to smokers and ex-smokers during the pandemic, as reported previously.

**Figure 3.**
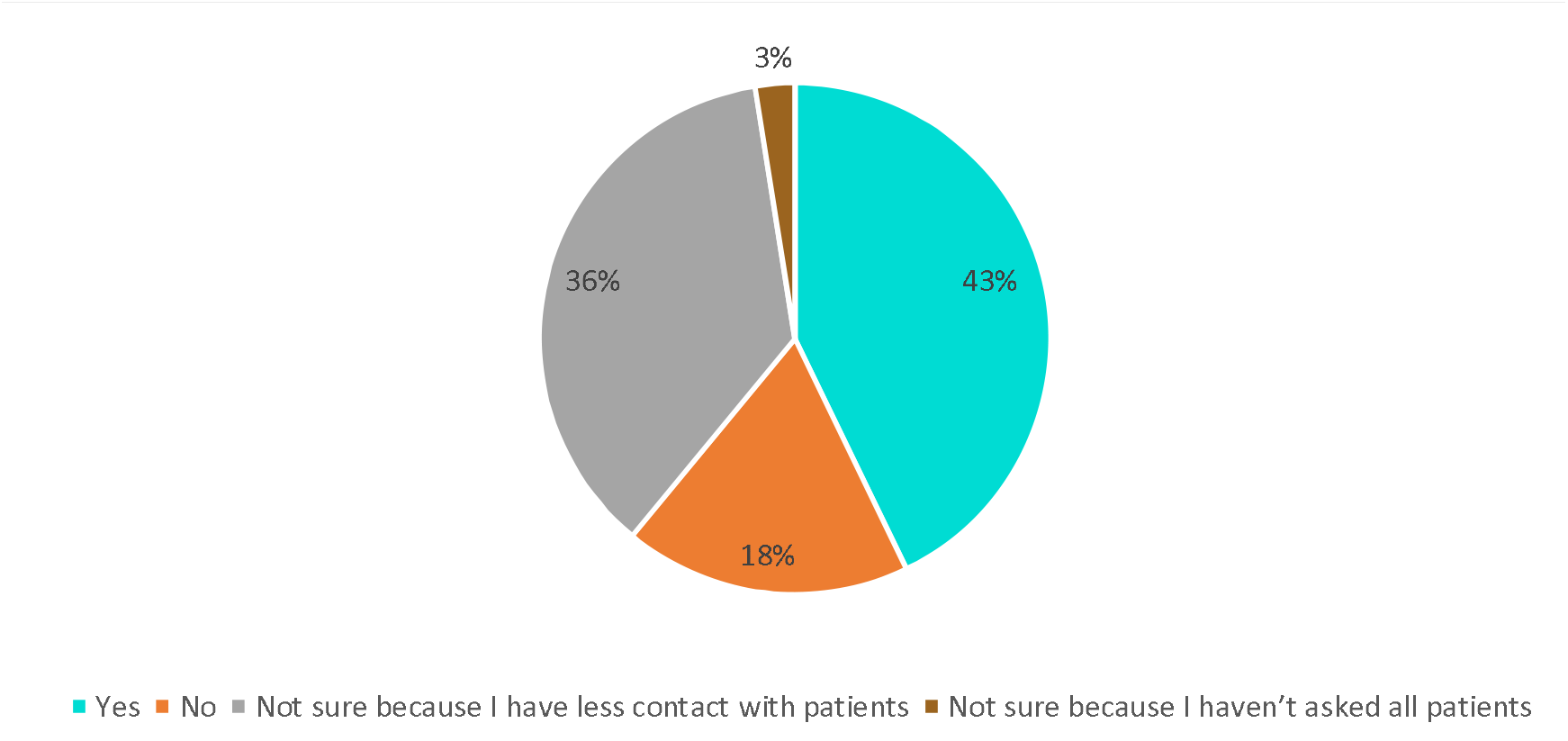
HCP perception of increased patient smoking during lockdown Question: Have you seen increased smoking from your patients since the UK’s lockdown?, N=677

### Self-reported changes in consumer smoking behaviours due to COVID-19

The following results pertain to the question ‘have there been any changes to your smoking behaviour as a result of the COVID-19 lockdown?’. 546 dual users and current smokers were asked this question; however, 57 respondents were excluded due to contradictory selections which prevented meaningful analysis (no change + more/less frequent, no change + more/fewer cigarettes, more frequent + less frequent, more cigarettes + less cigarettes, ‘other’). Multiple responses to the question were permitted, and results have been analysed in two different ways – Table 4 treats key response combinations as separate variables, while Figures 5–9 count the total number of responses.

**Table 4:** Changes in smoking behaviours as a result of the COVID-19 lockdown Question: Have there been any changes to your smoking behaviour as a result of the COVID-19 lockdown? (select all that apply)

| Response | Count of respondents | Percentage | Grouping | Count of respondents | Percentage |
| --- | --- | --- | --- | --- | --- |
| Smoking more frequently | 53 | 11% | Increased smoking behaviours | 146 | 30% |
| Smoking more cigarettes | 66 | 13% |  |  |  |
| Smoking more frequently + smoking more cigarettes | 27 | 6% |  |  |  |
| Smoking less frequently | 71 | 15% | Decreased smoking behaviours | 134 | 27% |
| Smoking fewer cigarettes | 45 | 9% |  |  |  |
| Smoking less frequently + smoking fewer cigarettes | 18 | 4% |  |  |  |
| No change | 163 | 33% | No change | 163 | 33% |
| Smoking less frequently + smoking more cigarettes | 23 | 5% | Changed smoking behaviours (neither increase nor decrease) | 27 | 6% |
| Smoking more frequently + smoking fewer cigarettes | 4 | 1% |  |  |  |
| Quit smoking | 14 | 3% | Quit smoking | 19 | 4% |
| Quit smoking + smoking more frequently | 0 | 0% |  |  |  |
| Quit smoking + smoking more cigarettes | 1 | 0% |  |  |  |
| Quit smoking + smoking less frequently | 2 | 0% |  |  |  |
| Quit smoking + smoking fewer cigarettes | 1 | 0% |  |  |  |
| Quit smoking + smoking fewer cigarettes + smoking less frequently | 1 | 0% |  |  |  |
| **TOTAL** | **489** | **100%** |  | **489** | **100%** |

Most consumers (67%) reported some kind of change to their smoking behaviour during the lockdown, however, there is not a clear trend as to whether more increased or decreased their smoking behaviours (Table 4). However, while 27% reported a decrease in their smoking, only 4% of respondents decided to quit smoking during the lockdown.

When comparing the frequency of individual responses between social grades, respondents in Grades A-C1 had higher proportions of respondents (27%-30%) who reported smoking more cigarettes overall than those in lower grades (10%-25%; Figure 4). The majority of respondents in Grades D & E reported no change in their smoking behaviour (52% and 59%, respectively) (Figure 4).

**Figure 4.**
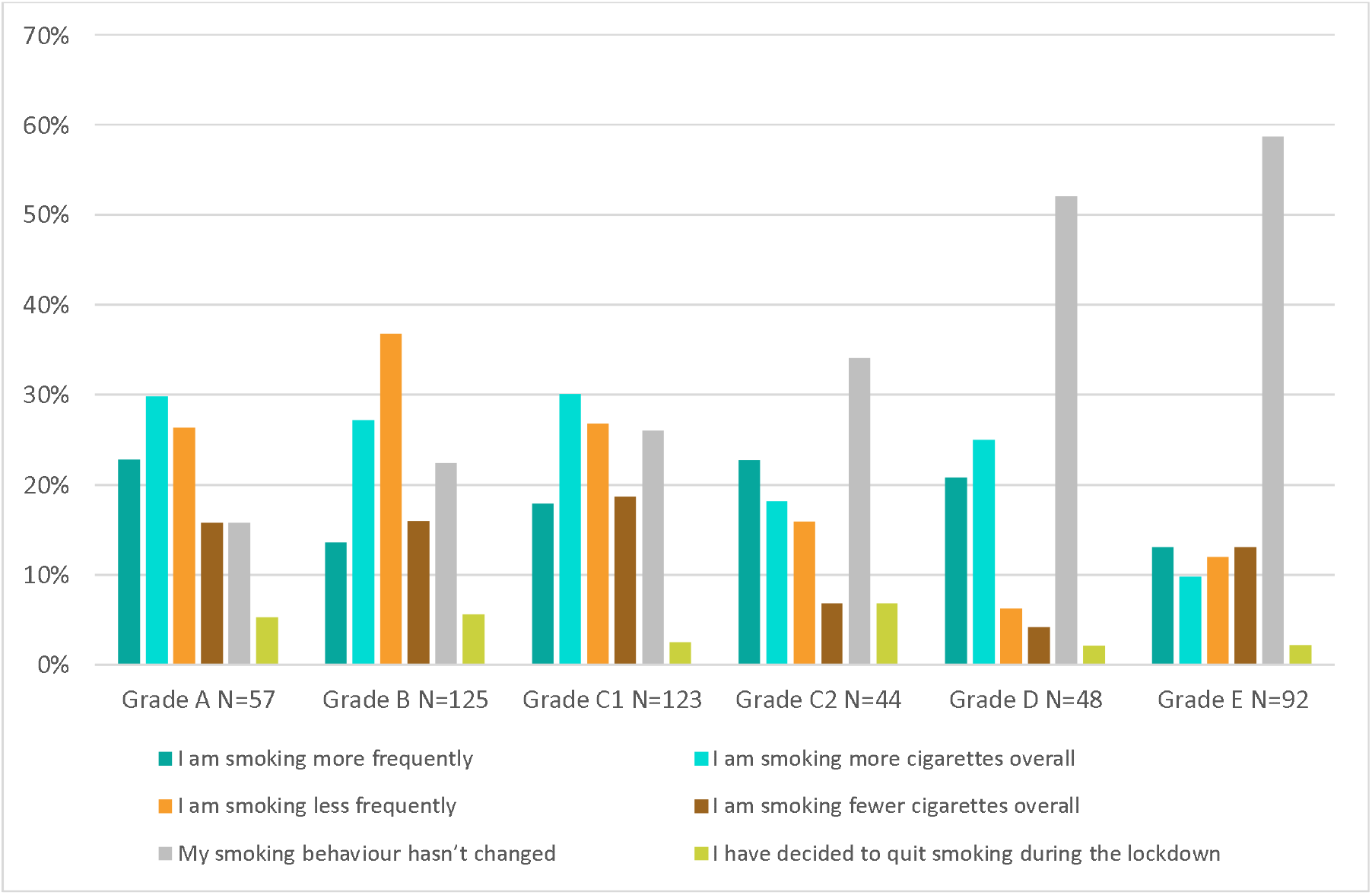
Changes in smoking behaviours as a result of the COVID-19 lockdown - comparing social grades Question: Have there been any changes to your smoking behaviour as a result of the COVID-19 lockdown? (select all that apply), N=489 respondents, 567 responses.

A greater proportion of dual users reported an increase in the number of cigarettes smoked during lockdown (27%) than current smokers (22%) (Figure 5). However, 34% of dual users reported smoking less frequently during the lockdown, compared to just 16% of current smokers (Figure 5).

**Figure 5.**
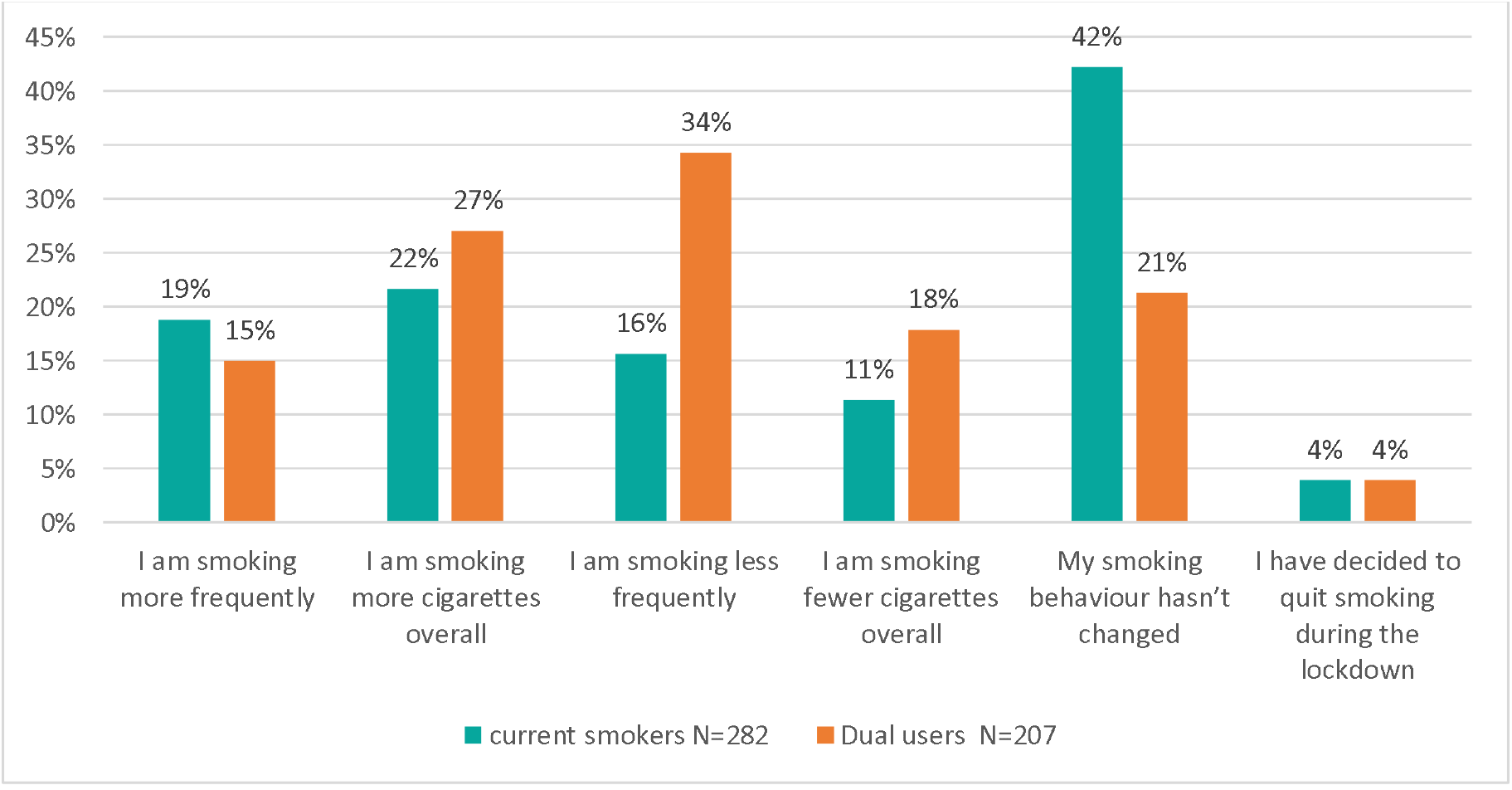
Changes in smoking behaviours as a result of the COVID-19 lockdown - comparing current smokers and dual users Question: Have there been any changes to your smoking behaviour as a result of the COVID-19 lockdown? (select all that apply), N=489.

A greater proportion of consumers aged 25-44 (36%-43%) reported smoking more cigarettes in lockdown than other age groups (14%-20%; Figure 6). Total 47% of consumers aged 45-74 reported no change in their smoking habits in lockdown compared to just 18% of 18-44 year olds (Figure 6).

**Figure 6.**
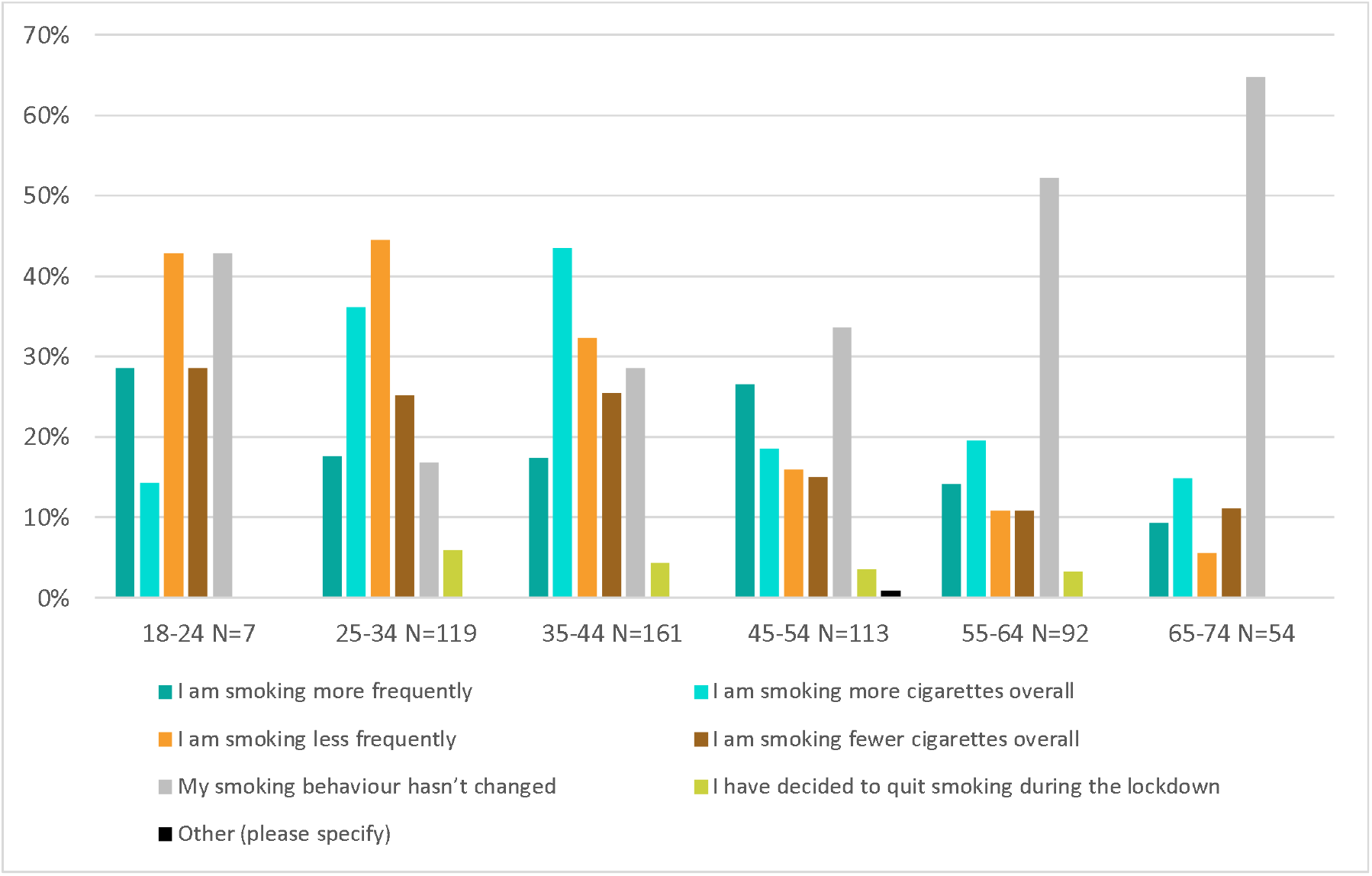
Changes in smoking behaviours as a result of the COVID-19 lockdown - comparing age groups Consumer survey Question: Have there been any changes to your smoking behaviour as a result of the COVID-19 lockdown? (select all that apply), N=546.

Consumers who reported smoking more frequently, and/or smoking more cigarettes overall during lockdown were asked to share the reason for this increase. The most commonly cited reason overall was boredom (66%), followed by stress (56%) (Figure 7). However, a greater proportion of dual users cited opportunity/freedom to smoke than current smokers (44% compared to 27%).

**Figure 7.**
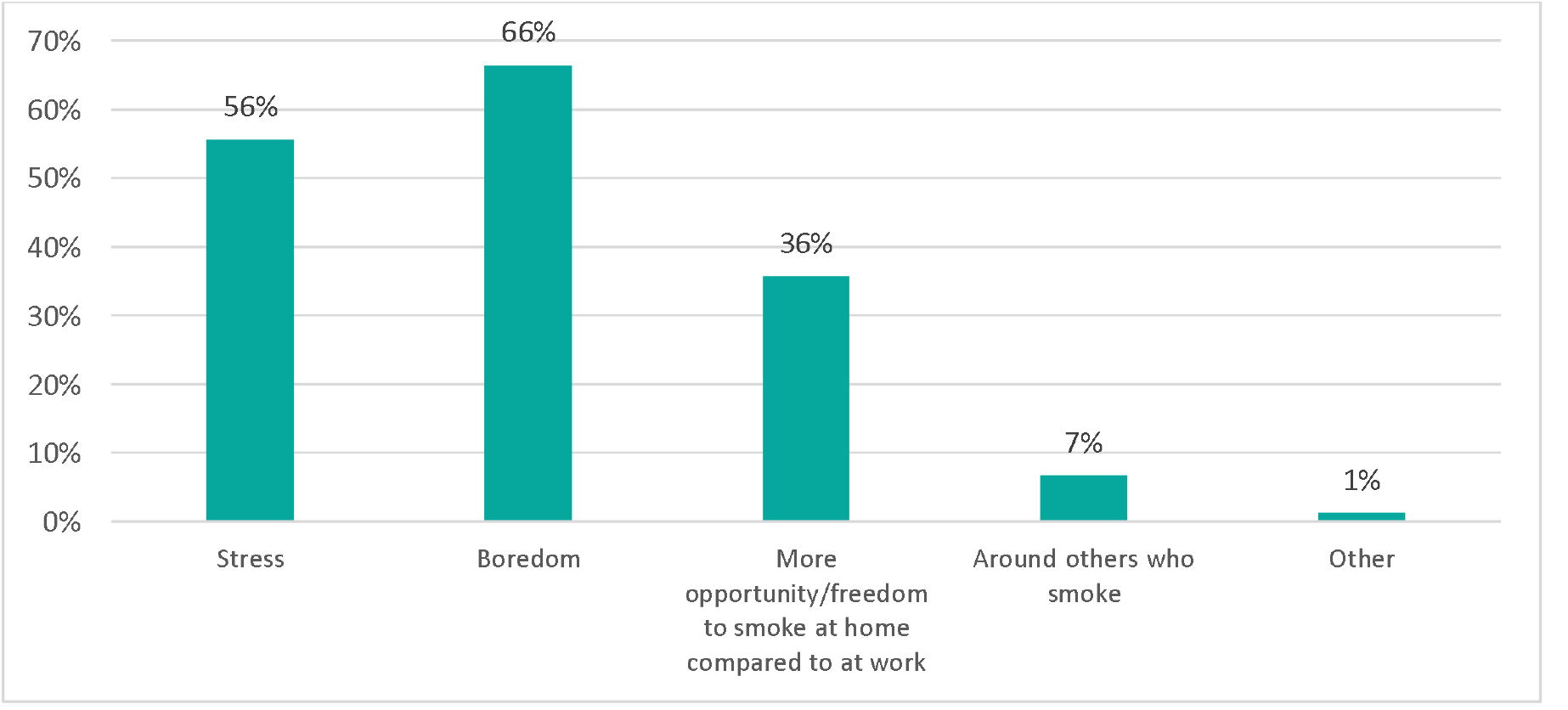
Reasons given for smoking more frequently and/or smoking more cigarettes during the COVID-19 lockdown Question: What is the reason for the increase in smoking? (select all that apply), N=241 respondents, 399 responses.

Greater proportions of respondents in social grades C2 (67%), D (75%), and E (79%) selected stress as the reason for their increase in smoking than those in higher social grades (grades A [61%], B [43%], and C1 [52%]; Figure 8).

**Figure 8.**
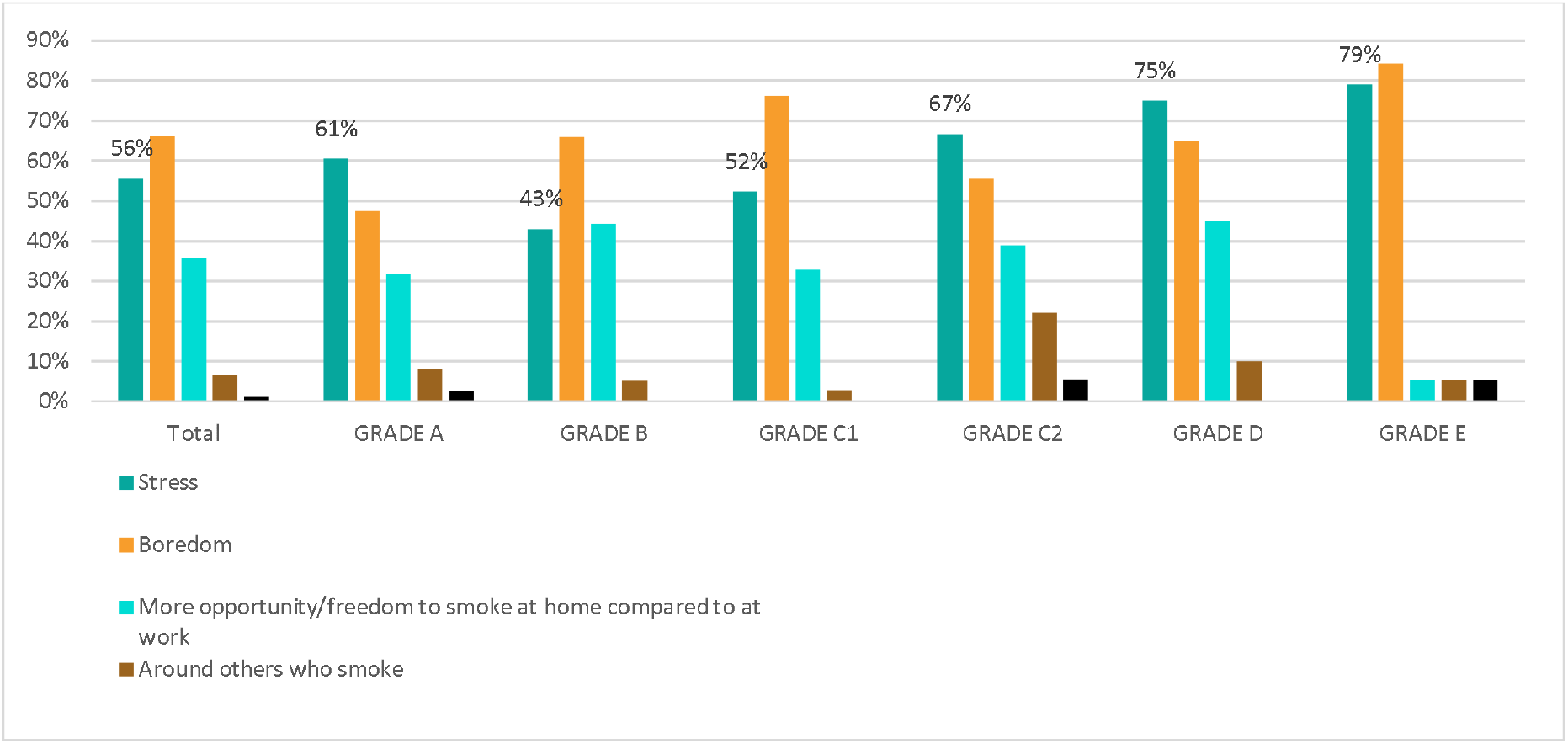
Reasons given for smoking more frequently and/or smoking more cigarettes during the COVID-19 lockdown - comparing social grades Question: What is the reason for the increase in smoking? (select all that apply), N=241 respondents, 399 responses.

### Impact of COVID-19 on consumer quit intentions

Overall, 36% of consumers stated that the COVID-19 crisis changed their plans on whether to quit smoking (a sample of 546 consumers comprised of dual users and current smokers). However, only 11 (6%) of consumers who changed their plans as a result of the COVID-19 crisis (n=195) also decided to quit during the pandemic.

### Impact of COVID-19 on the temptation to relapse

Of the 408 ex-smokers including ex-smokers who now vape, 15% said that they had been more tempted to start smoking cigarettes again during the pandemic. 55% of those in Grade A (n=20) reported being more tempted to start smoking again during the pandemic, the highest proportion of any social grade, followed by Grade B (n=73) at 27%. Of Grades C1 (n=114), C2 (n=57), D (n=39) and E (n=105), 15%, 9%, 5% and 8%, respectively agreed. 31% of 18-44 year olds (n=150) agreed that they had been more tempted to start smoking cigarettes again since the pandemic compared to just 6% of 45-74 year olds (n=258).

### Self-reported changes in consumer vaping behaviours due to COVID-19

The following results pertain to the question ‘Have there been any changes to your vaping behaviour as a result of the COVID-19 lockdown?’. Multiple responses to the question were permitted. Thus, the total number of responses counted in Figure 9 is higher than the number of respondents. In total 445 respondents were asked this question, however, 39 respondents were omitted from analysis due to contradictory responses (such as vaping more + less frequently)

**Figure 9.**
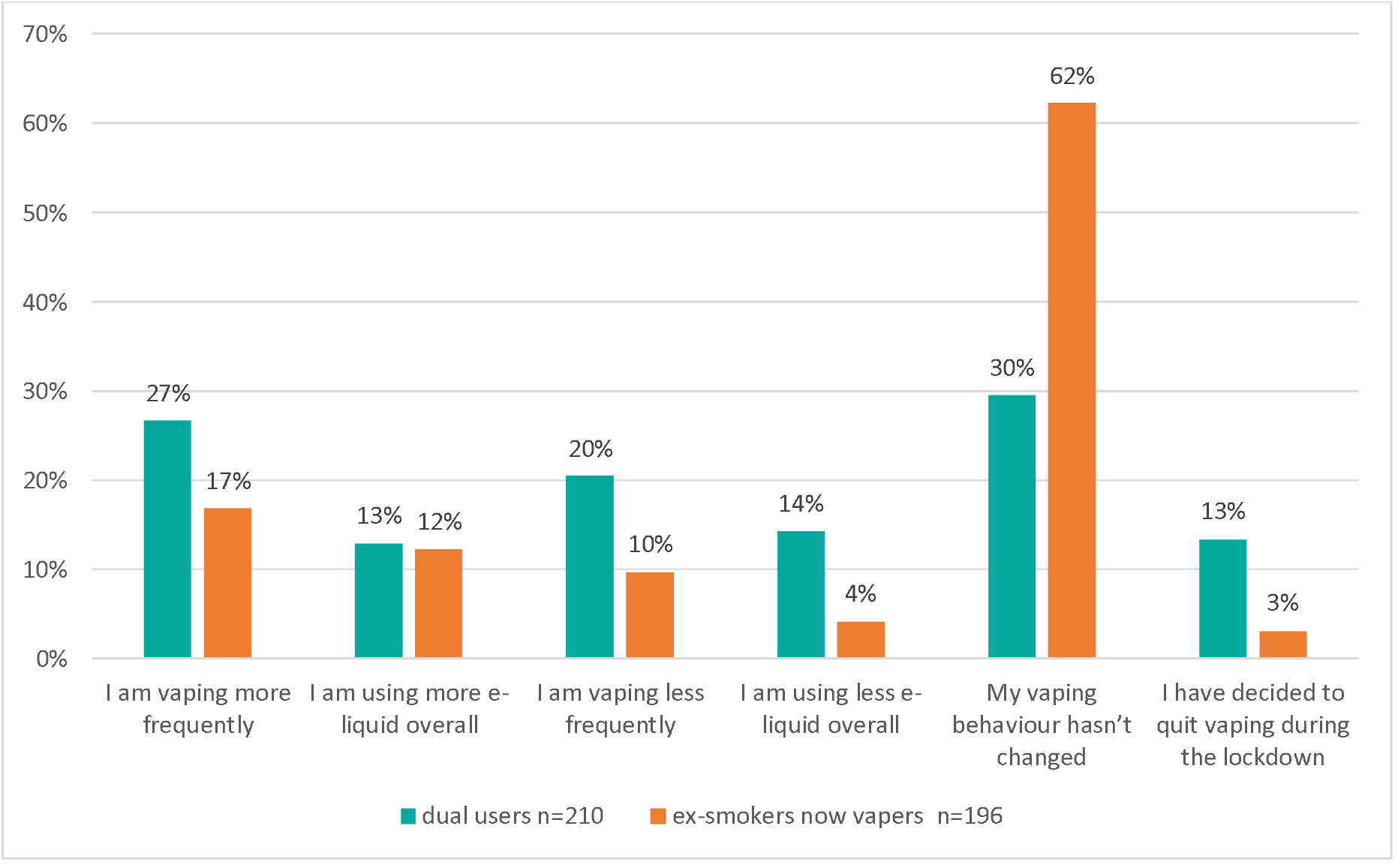
Changes in vaping behaviours as a result of the COVID-19 lockdown - comparing dual users and ex-smokers who vape Q. Have there been any changes to your vaping behaviour as a result of the COVID-19 lockdown?, N=406 respondents, 458 responses.

Higher proportions of dual users were vaping more frequently than ex-smokers who now vape (27% vs 17%) (Figure 9). Total of 62% of ex-smokers who vape reported no change to their vaping behaviours during the lockdown, compared to just 30% of dual users (Figure 9). A greater proportion of dual users decided to quit vaping during lockdown (13%) than ex-smokers who now vape (3%) (Figure 9).

Of the dual users who decided to quit vaping during lockdown (n=39), only 13% (n=5) also decided to quit smoking during this period.

Despite the closure of brick and mortar vape shops, the majority of vapers were able to access their preferred e-cigarettes and e-liquids during the pandemic (81%, n=445).

## Discussion

The UK’s journey to becoming smokefree (defined as <5% smoking prevalence) can be considered in its final laps, with a current smoking prevalence of 14%. However, it must not be forgotten that there are specific sub-populations and socio-economic groups that continue to have a higher-than-average smoking prevalence. This includes people in manual occupations, adults who are unemployed and those with a diagnosed mental health condition (8).

Our findings suggest that the government and the society’s response to COVID-19, resulting in changed workplace dynamics and healthcare delivery, may be having an unintentional but substantial impact on the behaviours of consumers and the support provided by their influencers. How this may affect long term smoking rates in the UK depends on how consumers and influencers are able to adapt to the “new normal”, as it begins to take shape.

Here we discuss key COVID-19 related observations from the survey and recommend interventions to address these.

While the vast majority of HCPs believed stressful situations, such as the COVID-19 pandemic, are likely to increase smoking and relapse, they do not appear to be consistently taking steps to address this risk. This suggests an obvious awareness-action gap. HCPs (excluding SSAs) proactively asked and documented patient smoking status during less than half of their interactions during the pandemic. Furthermore, a significant proportion of HCPs were unsure of whether they had seen an increase in smoking from their patients since the UK’s lockdown. With the exception of dentists and those shielding/unable to work, the HCP professions surveyed were largely expected to continue practising throughout lockdown, albeit with a shift to include significant proportion of virtual/telephone consultations. However, the scale and speed at which HCPs needed to adapt to remote consultations was unprecedented and may have been a totally new experience for many. Our findings suggest that HCPs, and their patients, are facing significant barriers to addressing smoking cessation during the lockdown. This may be due to HCPs focussing their energy on the primary reason for a consultation and on reducing Covid-19 related direct risks, as opposed to discussing smoking cessation as part of providing holistic care.

HCPs may benefit from training and support on how to effectively and efficiently address and prioritise smoking cessation during remote consultations. In identifying suitable support, learnings from practice nurses may prove valuable as they documented smoking status most frequently. It is important that with health on the top of the public agenda with the pandemic, established issues such as smoking are not forgotten, and instead are addressed as vital. It should be ensured that HCPs have the knowledge and means to enable them to document smoking status, as well as provide appropriate smoking cessation support as per their professional role, in all appropriate patient interactions.

Two-thirds of consumers reported a change in their smoking behaviour as a result of the COVID-19 lockdown. This is clearly a pivotal time, where new smoking habits can take hold and the impact of lockdown on smokers cannot be underestimated. In particular, adult smokers under 45 appear to be more at risk of changing their smoking habits and increasing the number of cigarettes smoked than those 45 and over. This age group, as a generally healthier age group, might not contact HCPs that often and hence might not get opportunistic advice from them. Furthermore, an increase in smoking among younger smokers can have a significant and long-term impact on the UK’s public health.

There is a need to take action to ensure that smokers receive appropriate support and that this time of change is used to address smoking and prevent it from undoing years of hard work from policymakers, HCPs and smokers themselves. Tailored interventions for different age groups may be beneficial. We also recommend that HCPs are made aware of this issue so they can support these people proactively, for example, by sending information on self-help smoking cessation tools and offering direct support via smoking cessation services.

Stress and boredom were consistently the top reasons given for an increase in smoking behaviour during the pandemic, and the strain it inflicts should not be underestimated. This was also recently documented in a clinical study from the United States (6). The uncertainty of the situation, resulting in changes to daily life, job security concerns, fear of the health impacts of COVID-19, and much more may have played a role. Healthier activities, that are safe to do in the circumstances, including music and sports, could be explored and promoted if suitable to help manage stress and boredom as well as directing smokers to appropriate counselling and support. Developing and increasing access to innovative smokefree apps that can address the behavioural component of smoking may play an important role here. Peer support groups, while face to face meetings are difficult, can be available on social media and the information of these on should be promoted more on reliable national smoking cessation websites like NHS smokefree.

Greater proportions of respondents in social grades A and B (managerial and professional occupations) reported changes in their smoking and vaping behaviours. Ex-smokers in social grades A, B and C1 also reported being more tempted to start smoking again during the pandemic, compared to those in grades C2, D and E. These differences between social grades may reflect the fact that those in social grades A, B and C1 are more likely to have experienced a transition to working from home during the pandemic, which may provide more opportunity to smoke than in the workplace while also exposing them to different social influences, along with stress and boredom. Furthermore, this has been a significant change in routine which is commonly associated with behavioural habits forming. This resurgence of smoking among the under-45 year old professionals is concerning. They are otherwise healthy, rarely seek general practice appointments or other health advice, and if not proactively given support to quit smoking now, could directly present with a tobacco related disease in the near future.

The impact of such significant changes to daily life as seen during the pandemic should not be underestimated and it is important to ensure that smokers and ex-smokers alike who have experienced significant changes have appropriate support. Beyond policymakers and HCPs, employers could play an important role in supporting employees who have experienced a change in work circumstances (whether that be working from home, furlough or redundancy) to direct them to appropriate smoking cessation support such as apps and local services.

Around two-fifths of respondents (36%) reported that they had changed their plans to quit smoking as a result of COVID-19. However, of those who changed their quit plans, only 6% decided to quit during the lockdown. This is very concerning as it suggests many smokers may have cancelled or postponed their quit plans during the pandemic. This may be due to multiple reasons including the wrong assumption that stop smoking services might not be functioning in lockdown, and not having enough knowledge about smoking cessation tools to access those on their own. There could also have been concerns around accessing quit smoking aids as COVID-19 fears and government advice may have made them reluctant or unable to visit shops. Another potential reason is the challenge of resisting smoking when spending so much time at home, as well as increased boredom and stress making quitting particularly difficult and smoking particularly tempting (as seen previously). However, as this is such an unusual but extended time there is a real risk that smokers may postpone their quit plans indefinitely as their behaviours during the pandemic become more entrenched.

More needs to be done to support smokers who intended to quit to follow through during the pandemic. A wider range of smoking cessation products and services should be available for consumers to choose from. In the case of the UK, this could include apps, regulated nicotine replacement products such as nicotine gums and e-cigarettes and in-person as well as online support from Stop Smoking Services. Influencers can play an important role here in being better informed about the cessation products and then advising consumers opportunistically on quitting smoking and on preventing relapse.

Overall, 4% of smokers surveyed decided to quit smoking during the lockdown, however, 27% reported a decrease in their smoking behaviours suggesting that some smokers have decreased their smoking during lockdown without having decided to quit. There are several potential reasons for this including:

- Smokers may have previously decided to quit, and are progressing along with their quit plan throughout the lockdown
- Smokers may have decided to reduce their smoking, instead of quitting fully during the pandemic, potentially due to concerns for their health and perhaps a view that reduction will offer certain protective benefits
- Smokers may no longer be in the environments and situations where they typically smoked, for example, they will not have been able to visit bars and nightclubs for the duration
- Smokers spending more time at home may be encouraged or pressured by other members of the household not to smoke as frequently

This reduction in consumption may be a valuable step towards stopping smoking long term.

Interventions that assist people with quitting during and after the pandemic are required, especially to support those who may be decreasing their smoking but not quitting. Circumstances are so different and are clearly impacting people’s quit decisions, therefore it should not be assumed that pre-COVID-19 practices are still as appropriate and effective in these circumstances. Interventions that are adapted to the uncertainty and changed behaviours of the pandemic are required urgently to avoid good progress from being lost and encourage smokers to use this time of change to quit. These interventions need to take into consideration multiple possible constraints such as another lockdown, reduced access to HCPs and vape shops, economic uncertainty, and the emotional toll of the pandemic.

Greater proportions of dual users reported increasing their cigarette consumption during lockdown than current smokers. Dual users were also more likely to state that the reason for this increase was due to more freedom/opportunity to smoke at home than at work, than current smokers. However, while almost double the proportion of dual users (34%) reported smoking less frequently during lockdown (compared to just 16% of current smokers), the same proportion of both groups decided to quit smoking during the lockdown.

Dual users also appear to have changed their vaping habits during lockdown more than ex-smokers who vape. Of the dual users who decided to quit vaping during lockdown only 13% also decided to quit smoking. While the sample is too small to draw a meaningful conclusion, this may imply that some dual users are ‘giving up’ on vaping when they no longer have restricted use of cigarettes during lockdown at home.

These initial findings may suggest that vaping is a far more consistent habit in ex-smokers who vape than in dual users, and that for dual users the role of vaping may be to enable them to access nicotine in locations where smoking is prohibited - a barrier reduced for many when they spend more time at home. While the majority of vapers were able to access their preferred vaping materials during the lockdown, 19% faced challenges, which may also have contributed to these changes in consumption.

Further research is warranted to understand what proportion of dual users have started vaping with the intention of quitting smoking permanently, and what proportion would prefer to revert to smoking exclusively if their environment permitted it. Such research may have broader implications on how e-cigarettes are sold, regulated and marketed in the UK. However, during COVID-19 dual users appear to be a group that would benefit from specific interventions that encourage them to continue with vaping as a means of avoiding cigarette consumption, and not to revert to smoking.

15% (around one in six) ex-smokers and ex-smokers who now vape reported being more tempted to start smoking cigarettes again since the pandemic. While this is not necessarily a representative sample, when we consider that there are an estimated 11.8 million ex-smokers in England alone (Public Health England, 2018) this could mean 1.8 million ex-smokers are at risk of returning to smoking. Relapse appeared to be more of a risk for ex-smokers who are younger (31% of 18-44 year olds were more tempted) and/or in higher social grades (55% of Grade A and 27% of Grade B were more tempted) highlighting the need for specific interventions to identify and support those groups most at risk.

What is also concerning is that when this group was asked what they were doing to resist relapse, none mentioned using typical stop smoking aids, or seeking professional help.

While only 26 responses were gathered, this could suggest that either ex-smokers are unaware of the support available, don’t believe they are ‘eligible’ for support as they are no longer smokers, or are facing other barriers. It is vital that the hard work is not lost and that ex-smokers continue to be supported – work needs to be done to clearly highlight the availability and accessibility of stop smoking aids during the pandemic. Furthermore, the language used in quitting may suggest a certain finality when people have quit, which while giving a sense of achievement may act as a barrier to seeking help again in the future.

If ex-smokers at risk of relapse can be supported early, this will likely be far more efficient and effective than waiting for them to regaining their status as smokers before seeking and offering help. There is a need for effective ways of proactively identifying ex-smokers at risk of relapse, and interventions that can support them before they start again, not once they relapse.

This research needs to be put into context of available smoking cessation tools and services. It will be crucial for innovators of behavioural tools for cessation (e.g. smartphone apps) and manufacturers of suitably regulated safer nicotine products to provide the necessary breadth of choice and sustainable way out of smoking. (12, 13)

### Strengths and limitations

This survey benefits from gaining perspectives from both HCPs and SSAs as well as a diverse range of smokers, vapers and dual users, giving a more comprehensive view of the topic than any one group alone.

A limitation of the study is the sample size and the sampling technique. The sample size in this study was not representative of the population. The analyses were descriptive and the study was not powered for statistical analysis.

Another limitation of the study is the fact that behaviours were self-reported, and respondents were asked to explain certain behaviours. Therefore, their responses may not reflect subconscious factors which may drive certain behaviours. Respondents may also show some social desirability bias and demand characteristics, although the fact that the surveys were taken individually online may have helped to mitigate these issues.

The surveys were also unable to take into consideration the emotional state of participants as they were responding, such as the time of day and whether influencers were pre or post-shift, as well as the impact of the COVID-19 crisis itself. Depending on their responses, some respondents will have completed surveys that are longer than is typically recommended, and therefore may have begun to experience fatigue, however, influencers in particular, were well remunerated due to being a specialist sample. The order of the questions in each survey attempted to follow a logical flow in order to make the surveys less mentally taxing to complete, however, this adds the risk that respondents may have been influenced by previous questions.

The classification of the social grade was entirely based on occupation and did not take into consideration other factors such as household income, or educational attainment and should be viewed critically. Furthermore, consumer and influencer groups were categorised by self-defined responses, and it cannot be guaranteed that the most appropriate options were selected by respondents in each instance.

## Conclusions

This research among current and former smokers and their influencers highlights the change in their behaviour during the COVID-19 pandemic. The unintended consequences of COVID-19 related responses by the government, employers and the public may derail the UK’s progress towards becoming smokefree. COVID-19 and its related social and health impact may linger for the foreseeable future. The recent report by Royal College of Physicians ‘Smoking and health 2021 A: coming of age for tobacco control?’ stated that in 2020, 80,000 people died of COVID-19 while 94,000 died of smoking related diseases in the UK. (10) This reiterates the scale of the negative impact smoking has on people’s mortality.’

This research underscores the need to keep smoking cessation and relapse prevention top on the healthcare agenda, with adjustments made to reflect the “new normal” reality of health-seeking behaviour and remote clinical practice. It is crucial to continue to understand the evolving challenges for the remaining smokers in the UK to quit smoking long term and address those challenges head-on. This includes (i) identifying the most reliable and trustworthy sources of guidance among the influencer networks and (ii) equipping these influencers with accurate information and necessary tools to help smokers on their quitting journey.

## Data Availability

The data related to this project is available on the website: www.smoke-free.uk

## Acknowledgements

CHRE were supported by Ogilvy Consulting’s Behavioural Science Practice in the design of the questionnaires and data analysis, and Atomik Research for executing the market research.

## Ethics approval

The individuals who were assessed in this survey had consented to be contacted for further research and were not recruited from National Health Service organisations. It was confirmed with the Health Research Authority that HRA and HCRW Approval was not required for this study (Communication dated 1st May 2020).

